# Anatomical Accuracy of Generative AI for Congenital Heart Disease Illustrations: Gemini NanoBanana Versus ChatGPT Models in a Blinded Comparative Study

**DOI:** 10.64898/2026.02.21.26346792

**Authors:** Abdullah Alhuzaimi, Abdulrahman Alkanhal, Ashwag Alruwaili, Nasser S. Alharbi, Fahad Alfares, Raniah N. Aldekhyyel, Samar Binkheder, Abdulrahman Temsah, Fadi Aljamaan, Muhammad Shahzad, Abdulilah Z Albriek, Frhan I. Alanazi, Duaa Abdulaziz Alhindi, Serin M. Al-khatib, Abdallah A. Darweesh, Ibrheem Altamimi, Amr Jamal, Khaled Saad, Khalid Hasan, Ayman Al-Eyadhy, Khalid H Malki, Mohamad-Hani Temsah

**Affiliations:** Department of Cardiac Sciences, College of Medicine, King Saud University, Riyadh, Saudi Arabia; Department of Cardiac Science, King Saud University Medical City, Riyadh, Saudi Arabia; Radiological Sciences Department, College of Applied Medical Sciences, King Saud University, Riyadh, Saudi Arabia; Pediatric Department, College of Medicine, King Saud University, Riyadh, Saudi Arabia; Medical Informatics and e-learning Unit, Medical Education Department, King Saud University, Riyadh, Saudi Arabia; College of Engineering and Advance Computing, Alfaisal University, Riyadh, Saudi Arabia; Department of Critical Care, King Saud University, Riyadh, Saudi Arabia; Pediatric Cardiac Intensive Care Unit, King Faisal Specialist Hospital & Research Center, Riyadh, Saudi Arabia; Cardiothoracic Imaging, Medical Imaging Dept, King Saud Medical City: Riyadh, Saudi Arabia; Department of Basic Sciences, Prince Sultan bin Abdulaziz College for Emergency Medical Services, King Saud University, Riyadh, Kingdom of Saudi Arabia; College of Medicine, Jordan University of Science and Technology, Irbid, Jordan; Department of Family Medicine, King Saud University, Riyadh, Saudi Arabia; The digital health clinical trials center, Seha Virtual Hospital, Ministry of Health, Riyadh, Saudi Arabia; Evidence Based Health Care & Translational Research Chair, Department of Family Medicine, King Saud University, Riyadh, Saudi Arabia; Pediatric Department, Faculty of Medicine, Assiut University, Assiut, Egypt; Organ Transplant Center of Excellence, King Faisal Specialist Hospital & Research Center, Kidney & Pancreas Health Center, Riyadh, Saudi Arabia; Research Chair of Voice, Swallowing, and Communication Disorders, Department of Otolaryngology-Head and Neck Surgery, College of Medicine, King Saud University, Riyadh, Saudi Arabia; College of Medicine, Alfaisal University, Riyadh, Saudi Arabia

**Author notes:** Correspondence: Mohamad-Hani Temsah, College of Medicine, King Saud University, Riyadh, Saudi Arabia. Equally contributed first authors.

**Keywords:** Generative artificial intelligence, text-to-image models, congenital heart disease, medical illustration, anatomical accuracy, educational utility, medical education, expert validation, image fidelity, comparative model performance

## Abstract

**Background:** Generative artificial intelligence (AI) systems are increasingly used to produce medical illustrations for education; however, their anatomical accuracy in complex domains such as congenital heart disease (CHD) remains insufficiently validated.

**Methods:** In an assessor-blinded comparative study, we evaluated AI-generated CHD illustrations from two contemporary text-to-image platforms (ChatGPT-5/ChatGPT-Images and Gemini NanoBanana) against human-modified educational images. Twenty different CHD types were included, yielding 147 images that were assessed by 20 physicians (10 CHD experts and 10 non-specialists). Images were rated across four domains: anatomical accuracy, label usefulness, visual attractiveness, and suitability for medical education (total score range, 4–12).

**Results:** Among 2,940 total image evaluations, the human-modified images demonstrated the highest anatomical accuracy (48.3% rated accurate), followed by NanoBanana (22.7%), while ChatGPT-generated images were predominantly rated as fabricated or incorrect (86.3% for ChatGPT-5 and 85.2% for ChatGPT-Images; p<0.001). Educational usability “as is” was highest for the human-modified images (37.9%) compared with NanoBanana (13.1%) and ChatGPT platforms (≤2.1%; p<0.001). Median overall quality scores were 8 for the human-modified CHD images and NanoBanana, versus 4 for both ChatGPT systems (p<0.001). In multivariable analysis, NanoBanana images were the closest to the human-modified images in quality (95% CI, 0.91–0.98), while ChatGPT-Images (95% CI, 0.58–0.63) and ChatGPT-5 (95% CI, 0.55–0.59) showed marked quality reductions.

**Conclusions:** The current generative AI systems produced visually compelling but frequently anatomically inaccurate CHD illustrations, falling substantially short of the current educational standards. Model choice strongly influences performance, with Gemini NanoBanana outperforming ChatGPT-based systems yet remaining inferior to expert-designed human-modified images. AI-generated cardiac imagery should be used only within expert-reviewed educational workflows rather than as independent instructional resources.

## Introduction

Anatomical images are crucial in medical education and enable learners to infer organ shape, size, and spatial relationships that cannot be directly observed in living patients. In a long-standing practice in medical education, these visuals were integrated across lectures, ,practical sessions, small-group teachings, atlases, and digital platforms, where they function as a shared visual medium for teaching, discussion, and clinical reasoning [1]. In congenital heart disease (CHD), the visual representations of three-dimensional relationships among chambers, valves, and great vessels, which are key to understanding the pathology and guiding medical and surgical interventions, are difficult to communicate with text alone. Evidence from CHD-focused educational interventions showed that adding tangible or advanced 3D representations, such as printed or modeled hearts, improved learner understanding and confidence Evidence from CHD-focused educational interventions further showed that adding tangible or advanced 3D representations, such as printed or modeled hearts, improved learner understanding and confidence [2]. Reinforcing the specific value of spatial visualization for CHD morphology. These high-quality anatomical illustrations were not merely descriptive; when intentionally designed, they enhanced learning efficiency by emphasizing diagnostically or conceptually salient features while suppressing irrelevant detail. This educational approach aligned with cognitive load theory proposed by John Sweller [3], an Australian educational psychologist and later developed by others [4], which suggested that instructional formats that minimized unnecessary processing demands could preserve limited working-memory capacity for schema construction and more durable learning. Complementary learning theories supported the same mechanism of combined visual and verbal input; consistent with dual coding theory, predicted improved retention and understanding when information was encoded through coordinated verbal and visual channels [5]. In practical terms, clear illustrations guided attention, reduced extraneous cognitive load, and helped learners construct accurate internal representations of anatomy that could be applied to clinical explanation and problem solving [1].

Anatomical cardiac images were typically obtained from textbooks and atlases, as well as cadaveric photographs. Clinical imaging modalities, including echocardiography, CT, and MRI, further expanded access to cardiac anatomy while remaining noninvasive for patients. Surgical photographs and curated clinical case images were also incorporated to enhance clinical relevance and contextual understanding. Educators increasingly relied on 3D software or 3D models to convey spatial relationships that 2D figures could not easily show [6]. Despite their value, traditional resources carried practical constraints. High-quality images were not always available for the exact lesion, view, or learning goal. Licensing atlas images or specialized 3D tools could be costly. Customization was often limited, particularly when educators needed the same anatomy shown in a consistent orientation across multiple slides. Even in standard resources, variation in labeling or left–right orientation could confuse learners and reduce learning effectiveness [7,8]. Licensing atlas images or specialized 3D tools is also costly [9].

Generative artificial intelligence (GenAI) was increasingly discussed and adopted across health professions education as a practical “workflow partner” for learners and faculty. It provided; a) rapid synthesis of complex content, b) assistance with academic writing, c) formative feedback and learning support that felt immediate and personalized [10]. In principle, AI image tools could provide great, and practical tailored CHD visuals on demand. However, from an anatomy education point pf view, this promise extended beyond text: AI text-to-image systems were positioned as fast and low-cost tools that could rapidly generate multiple visual alternatives and reduce some barriers [11]. This was created by licensing constraints and limited access to specialized illustration expertise but considering AI as pedagogically non-neutral.

However, GenAI suffered serious drawbacks in uncritical reliance which was linked to cognitive offloading, susceptibility to biased or incorrect outputs, and erosion of independent reasoning when tools were used without supervision or contextual understanding [10]. These concerns became even more consequential when GenAI outputs were used as visual teaching material, because confident-looking images could invite over trust while quietly embedding major structural errors that learners would readily internalize [12]. The human heart is an ideal stress test for generating scientific images because cardiovascular learning depends on spatial truth, not just visual plausibility. It is believed that learners must integrate standard views (anterior, posterior, inferior) to understand how chambers form surfaces and how coronary arteries course across them, often using 3D, cutaway, or transparent depictions. In this context, small errors in orientation or vessel trajectories can teach the wrong anatomy. This need for fidelity became sharper because the images can be visually compelling while still getting the anatomy wrong. In Noel’s assessment of text-to-image outputs, cardiac illustrations often missed correct coronary origins and distorted branching of the aorta and pulmonary trunk, the kind of subtle errors that can quietly recalibrate a learner’s mental model in the wrong direction [11]. At the institutional level, deploying generative AI in health professions education is less a question of picking a tool and more a question of governance. Effective adoption needs to be explicitly human-centered, with defined requirements for transparency and accountability, and with guardrails that align day-to-day use with institutional and national policy obligations [13]. UNESCO’s guidance emphasized that rapid tool release had outpaced regulatory readiness, leaving user data privacy insufficiently protected and intellectual property and copyright risks an added practical constraint [14]. Unless institutions and regulators mandated safeguards and responsible implementation.

Rapid enthusiasm for GenAI in anatomy education has surpassed the current evidence base on anatomical fidelity, which remains limited and, across studies, shows mixed and inconsistent accuracy. Early evaluations showed that text-to-image models could produce visually convincing anatomy. Yet, embedding non-trivial errors, including clinically meaningful inaccuracies in cardiac structures and vessel branching patterns [11]. When these errors were examined more closely, it became clear that they were not generic property of GenAI; they emerged depending on the model, its configuration, and the prompt-to-image pipeline. Consistent with this, side-by-side testing of multiple generators on the same core anatomy prompts, including the human heart, showed substantial platform-to-platform differences in correctness [15]. In aggregate, these findings justified the need for a focused, tool-comparative study of congenital heart disease image generation, providing the kind of evidence institutions and educators needed to make defensible, safety-conscious decisions about curricular integration.

Comparative data are limited on how different state-of-the-art and legacy systems performed when tasked with CHD imagery, and whether learners and clinicians judged the results as accurate, appealing, and educationally useful. Therefore, this study aimed to evaluate AI-generated CHD images produced with Gemini Nano Banana and to compare them with outputs from ChatGPT-5 and legacy DALLE-3. We assessed (1) accuracy, defined as anatomical and lesion-specific correctness, (2) Labels usefulness (3) appeal, defined as perceived clarity and visual quality, and (4) educational value, defined as perceived usefulness for learning and teaching CHD anatomy.

## Method

### Study Design and AI Image Generation Procedures

This assessor-blinded model evaluation study examined the anatomical accuracy, educational usefulness, and visual quality of AI-generated illustrations of CHDs using three contemporary text-to-image platforms: OpenAI’s ChatGPT-5, ChatGPT-Images, and Google’s Gemini NanoBanana. The study design and scoring framework were adapted from the authors’ previously published work evaluating AI-generated CHD illustrations [12].

Twenty common CHDs and the normal human heart were included. The 20 CHDs included were selected based on established relevance to pediatric cardiology teaching and clinical practice, as illustrated in our previous research [12]. The normal heart was added as a separate category to provide a baseline for evaluating model consistency and evaluator discrimination.

For each anatomical category, evaluators assessed eight images presented in blinded random order: three images generated by ChatGPT-5, three images generated by Gemini NanoBanana, one image generated by ChatGPT-Images (released on 16 December 2025), and one human-modified image derived from authoritative open-access anatomical sources. This yielded a total of 168 images (21 anatomical categories × 8 images per category) evaluated under identical conditions.

All AI-generated and AI-resembling reference images were produced between December 10 and December 20, 2025. During this period, the ChatGPT-Images model was released; therefore, one additional image per anatomical category was generated using this platform and incorporated into the evaluation set.

To ensure consistency and reflect typical educational use cases, all AI-generated images were required to include internal text labels, and prompts were standardized across all platforms. The prompt used for each CHD and the normal heart was:

“Draw an accurate medical illustration of [CHD / normal human heart] designed for medical students, with clear and correct text labels inside the image to clarify the anatomical structures.”

For each anatomical category, Gemini NanoBanana generated three images, ChatGPT-5 generated three images, ChatGPT-Images generated one image, and one human-modified comparator image was included. All 168 images were subsequently uploaded into the evaluation platform (Appendix 1).

### Preparation of the Human-modified AI-Rendered Images

To optimize blinding integrity, the original human-modified illustrations were not presented directly to evaluators. Instead, each reference illustration was rendered using a third AI platform not included in the comparative analysis, producing an AI-stylized version that preserved full anatomical correctness while harmonizing the visual appearance across all eight images within each anatomical category and removing stylistic cues that could reveal the gold-standard source. These AI-resembling reference images, hereafter termed “human-modified images,” served as the comparator condition in all subsequent analyses.

### Blinding and Randomization

Blinding and randomization were implemented through a secure online evaluation interface on SurveyMonkey platform, with each evaluator receiving a uniquely randomized presentation of images. For each anatomical category, the eight images were displayed in random order, standardized in general format and visual style, and shown without identification of their generating source or type. Evaluators were therefore unaware of which images were produced by each AI model and which image represented the human-modified reference standard. This structured blinding approach minimized source-identification bias and strengthened the internal validity of inter-model comparisons.

### Participants (Evaluators)

A total of 20 evaluators participated in the study, comprising 10 CHD experts, including pediatric cardiologists and pediatric consultants, and 10 non-CHD healthcare professionals representing clinical backgrounds that commonly utilize anatomical illustrations in practice, such as internal medicine, nursing, and allied health. This deliberate expert versus non-expert grouping was designed to assess differences in anatomical discernment and to examine potential susceptibility to visually appealing yet anatomically inaccurate AI-generated imagery.

### Image Evaluation Framework

Each image was independently evaluated using a four-domain assessment tool adapted from prior work. The domains included anatomical accuracy (rated as accurate, partially correct, or fabricated/incorrect), usefulness and correctness of text labels (useful, midway, or useless/incorrect), visual attractiveness to medical professionals (attractive, midway, or not attractive), and suitability for medical education (usable as-is, usable after modification, or not suitable). Each domain was scored on a three-point scale (3–1), and a Total Image Quality Score ranging from 4 to 12 was calculated by summing the four domain ratings for each image. In total, evaluators completed 3,360 independent image assessments (168 images × 20 evaluators).

### Statistical Analysis

Descriptive statistics, including means, standard deviations, and frequency distributions, were used to summarize scoring patterns across the four evaluation domains. Inferential analyses were conducted to compare performance across image sources and evaluator groups. Chi-square tests were used for categorical comparisons between expert and non-expert evaluators, while Mann–Whitney U and Kruskal–Wallis tests were applied for comparisons of ordinal scores. A mixed-effects generalized linear model was constructed to examine predictors of overall image quality, with rater ID specified as a random effect and image source (ChatGPT-5, ChatGPT-Images, NanoBanana, and human-modified reference) entered as fixed factors. CHD complexity and evaluator expertise were included as covariates. Spearman correlation coefficients were calculated to assess relationships among the four scoring domains. Statistical significance was defined as p < 0.05.

### Ethical Considerations

Ethical approval was obtained from the Institutional Review Board (IRB) at King Saud University (Approval #25/0942/IRB). No patient data, personal identifiers, or human-subject interventions were involved. The project consisted solely of internal academic evaluation of AI-generated illustrations. All procedures involving human participants were conducted in accordance with the ethical standards of the institutional research committee and with the Declaration of Helsinki.

## Results

The images of normal heart and twenty other cardiac anomalies were evaluated by twenty physicians with varying levels of cardiac expertise. Cardiology experts represented 25% of the sample (n = 5), and an additional 25% (n = 5) were pediatric consultants. The largest proportion of participants consisted of non-cardiology physicians, who accounted for 50% of the sample (n = 10).

Evaluators’ perceptions of the overall complexity of various cardiac anomalies are shown in Figure 1. When ranked in descending order of perceived complexity, several anomalies were overwhelmingly regarded as complex by most physicians, with highest ranking lesions of Pulmonary atresia with ventricular septal defect and major aortopulmonary collateral arteries (PA, VSD, and MAPCA’s) was perceived as the most complex condition, with 95% of respondents classifying it as complex. At the lower end of the complexity spectrum, simple left to right shunt lesions such as atrial septal defect (ASD), patent ductus arteriosus (PDA), and ventricular septal defect (VSD) were predominantly classified as simple, with 90% or more of respondents endorsing this classification

**Figure 1:**
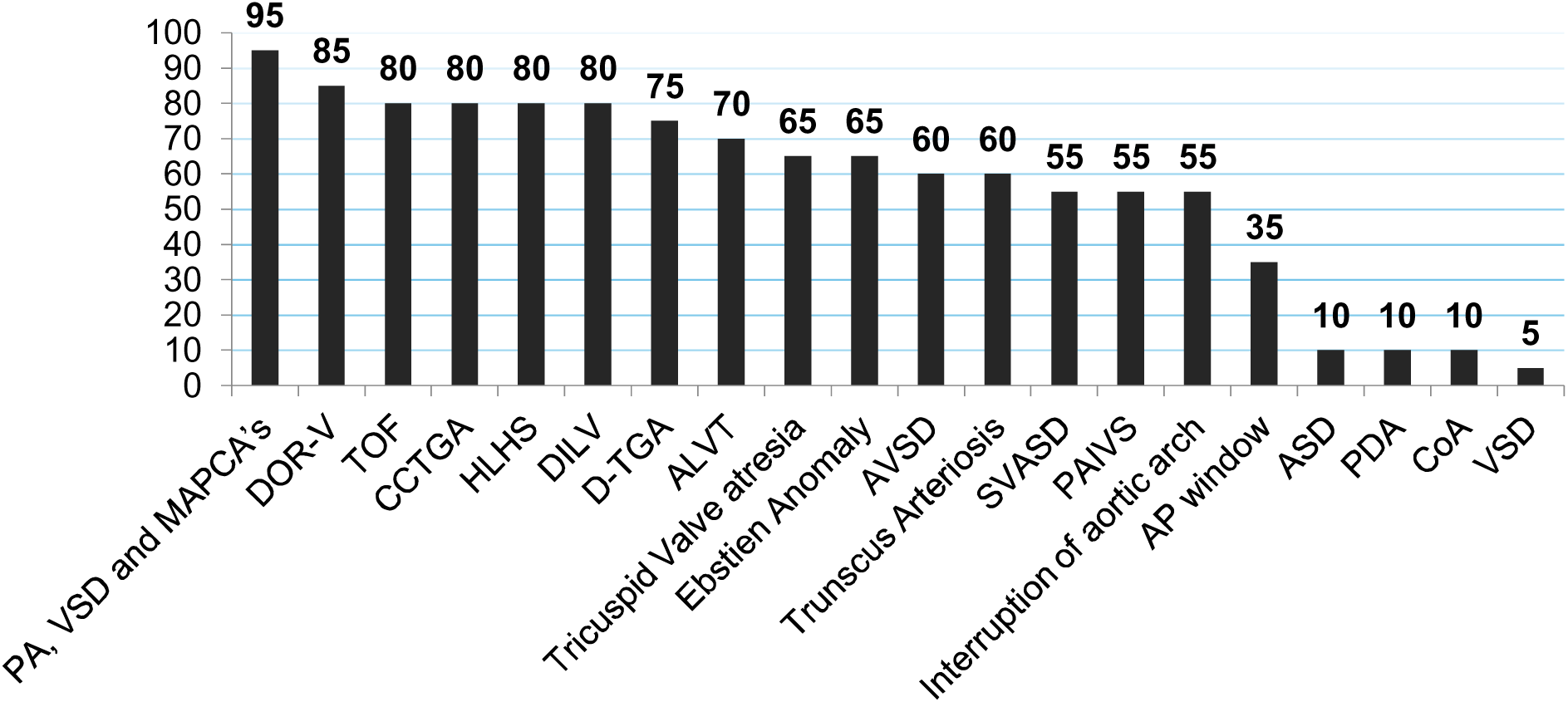
The Healthcare Workers Perceived Complexity of Cardiac Anomalies (complex %) Abbreviations: Pulmonary atresia with ventricular septal defect and major aortopulmonary collateral arteries (PA, VSD, and MAPCA’s), double outlet right ventricle (DORV), tetralogy of Fallot (TOF) , congenitally corrected transposition of the great arteries (CCTGA), hypoplastic left heart syndrome (HLHS), double inlet left ventricle (DILV), D-transposition of the great arteries (D-TGA), Aorto-Left Ventricular Tunnel (ALVT), atrioventricular septal defect (AVSD), sinus venosus atrial septal defect (SVASD), pulmonary atresia with intact ventricular septum (PAIVS), Aortopulmonary window (AP window), atrial septal defect (ASD), patent ductus arteriosus (PDA), coarctation of the aorta (CoA), ventricular septal defect (VSD)

### Evaluators’ ratings of AI-generated cardiac images from different Chatbots

The rating of images regarding the four different domains (anatomical accuracy, text usefulness, attractivity, usefulness for medical education) were compared among the different chatbots (ChatGPT, NanoBanana) and the human-modified images. Significant differences were observed across all evaluated dimensions, including anatomical accuracy, usefulness of text labels, visual attractiveness, usability for medical education, overall image quality, and time required for image generation (all p < .001).

With respect to anatomical accuracy (Table 1), the reference human-modified CHD Images demonstrated the highest proportion of images rated as accurate (48.3%), followed by the NanoBanana images (22.7%) and ChatGPT-5 2.8% and ChatGPT-Images 3.3% as accurate. On the other hand, most images generated by ChatGPT-5 and ChatGPT-Images were rated as fabricated (86.3% and 85.2%, respectively). The association between image source and anatomical accuracy was statistically significant, p < .001, indicating strong dependence of perceived anatomical accuracy on the generating source.

**Table 1:** Bivariate Analysis of Chatbot-Generated Cardiac Anomalies Image: Comparison Across Different Image Sources N = 3360 image ratings.

|  |  |  |  |  |  |
| --- | --- | --- | --- | --- | --- |
| **Category** | **Source of Image** |  |  |  | **p-value** |
|  | **ChatGPT-5** | **NanoBanana** | **human-modified images** | **ChatGPT-Images** |  |
| **Anatomical accuracy of image** |  |  |  |  | <0.001 |
| Accurate | 35 (2.8) | 286 (22.7) | 203 (48.3) | 14 (3.3) |  |
| Midway | 138 (11) | 503 (39.9) | 106 (25.2) | 48 (11.4) |  |
| Fabricated | 1087 (86.3) | 471 (37.4) | 111 (26.4) | 358 (85.2) |  |
| **Usefulness of text for the viewer** |  |  |  |  | <0.001 |
| Useful | 10 (0.8) | 308 (24.4) | 166 (39.5) | 11 (2.6) |  |
| Midway | 160 (12.7) | 574 (45.6) | 137 (32.6) | 73 (17.4) |  |
| Not useful | 1090 (86.5) | 378 (30) | 117 (27.9) | 336 (80) |  |
| **Attractivity of image for viewer** |  |  |  |  | <0.001 |
| Attractive | 39 (3.1) | 436 (34.6) | 69 (16.4) | 17 (4) |  |
| Midway | 233 (18.5) | 531 (42.1) | 160 (38.1) | 111 (26.4) |  |
| Not attractive | 988 (78.4) | 293 (23.3) | 191 (45.5) | 292 (69.5) |  |
| **Usability of image for medical education** |  |  |  |  | <0.001 |
| As is | 1 (0.1) | 165 (13.1) | 159 (37.9) | 9 (2.1) |  |
| after modification | 75 (6) | 600 (47.6) | 124 (29.5) | 34 (8.1) |  |
| Not useful for use | 1184 (94) | 495 (39.3) | 137 (32.6) | 377 (89.8) |  |
| **Overall Image quality rating (sum of four factors), median (IQR)** | 4 (1) | 8 (3) | 8 (6) | 4 (1) | <0.001 |
| **Time taken (seconds) for the chatbot to generate the images, median (IQR)** | 98 (30) | 31 (10) | NA | 70 (22) | <0.001 |

Regarding the usefulness of text labels, the human-modified CHD Images again achieved the highest proportion of ratings classified as useful (39.5%), followed by NanoBanana images (24.4%). In contrast, text annotations generated by ChatGPT-5 and ChatGPT-Images were predominantly rated as not useful (86.5% and 80%, respectively). The observed differences across sources were statistically significant, p < .001, underscoring marked variability in the educational value of embedded textual information.

In terms of visual attractiveness, NanoBanana images were most frequently rated as attractive (34.6%), outperforming human-modified CHD Images (16.4%) and both ChatGPT versions (3.1% for ChatGPT-5 and 4.0% for ChatGPT-Images). The human-modified CHD Images were more commonly rated as midway in attractiveness (38.1%), with p-value < 0.001.

Evaluation of usability for medical education revealed that Human-modified CHD Images were most frequently rated as usable “as is” (37.9%), followed by NanoBanana images (13.1%). In contrast, images generated by ChatGPT-5 and ChatGPT-Images were overwhelmingly rated as not useful for educational purposes (94.0% and 89.8%; respectively). NanoBanana images were most often perceived as usable after modification (47.6%), suggesting potential value as adaptable educational resources.

The overall image quality rating score (reflecting the sum of four domains rating) (accuracy, attractivity, usefulness of text, and usability of image for education ratings) and the resulted overall image quality score was measured with showed 6.22 points out of 12 points, SD=2.55 Points. Median overall quality scores were highest for NanoBanana and the human -made images (both median overall quality score = 8), whereas ChatGPT-5 and ChatGPT-Images demonstrated substantially and significantly lower median scores (both median overall quality score = 4), indicating inferior aggregated performance across quality domains.

### Comparison Of AI-Generated Cardiac Image Rating Across Different Physician Roles

The bivariate analysis (Table 1 supplementary) examining differences in image ratings across physicians with different clinical roles, including cardiology experts, pediatric consultants, and non-cardiology medical experts showed a statistically significant difference across all evaluated domains but the difference was small.

Non-cardiology medical experts reported a slightly higher median overall quality rating (median = 5, IQR = 4) compared with cardiology experts and pediatric consultants (both median = 4, IQR = 4), indicating a more favorable overall perception among non-specialists. Additionally, perceived cardiac anomaly complexity also varied significantly by clinical role, χ²(2) = 13.2, p = .001. Cardiology experts reported the highest median complexity ratings (median = 3, IQR = 1.25), followed by non-cardiology medical experts (median = 2.75, IQR = 1.25), whereas pediatric consultants reported the lowest median complexity (median = 2.38, IQR = 1.5). This finding suggests that specialized cardiac training is associated with heightened sensitivity to anatomical and clinical complexity.

### Multivariable Analysis of Evaluators Perceived Overall Image Quality Score AI Cardiac Images

Multivariable generalized linear model (GLM)was used to examine the factors associated with the overall quality of cardiac anomaly images rating score . The overall image quality score was calculated as the sum of the four evaluated dimensions. (Table 2)

**Table 2:** Multivariable Generalized Linear regression with Gamma for the images overall quality.

| Parameter | Multivariable adjusted Risk Rate | 95% CI for RR |  | p-value |
| --- | --- | --- | --- | --- |
|  |  | Lower | Upper |  |
| **(Intercept)** | 8.170 | 7.788 | 8.570 | <.001 |
| **Image source = ChatGPT-Images** | 0.600 | 0.577 | 0.625 | <.001 |
| **Image source =NanoBanana** | 0.943 | 0.913 | 0.975 | <.001 |
| **Image source =ChatGPT-5** | 0.569 | 0.551 | 0.588 | <0.001 |
| **Image source =Reference comparison - Open-source Human-modified CHD image** | 1 |  |  |  |
| **[Clinical Role of evaluators = Non-cardiologist medical experts** | 1.048 | 1.022 | 1.076 | <.001 |
| **[Clinical Role of evaluators=Pediatric consultants** | 0.977 | 0.947 | 1.008 | 0.147 |
| **Pediatric & adult Pediatric cardiologists** | 1.000 |  |  |  |
| **Mean Perceived cardiac anomalies complexity score** | 0.989 | 0.978 | 1.000 | 0.056 |
| *Dependent Variable: Overall cardiac anomalies Image Quality (Higher score denotes better usefulness, accuracy, usability and text usefulness). Estimation methodL Maximum Likelihood with Gamma Loglink.* |  |  |  |  |

After adjustment, image source emerged as the strongest predictor of overall image quality (Table 2). Compared with the reference category of human-modified open-source images, all AI-generated image sources were associated with significantly lower overall quality ratings. Images generated by the ChatGPT-Images demonstrated a 40% lower adjusted quality score (RR = 0.60, 95% CI [0.58, 0.63], p < .001), images from the ChatGPT-5 version (RR = 0.57, 95% CI [0.55, 0.59], p < .001). In contrast, NanoBanana-generated images exhibited modestly the smallest decrement in quality relative to human-modified images (RR = 0.94, 95% CI [0.91, 0.98], p < .001), indicating performance that approached but did not fully match human-modified open-source images.

The clinical role of evaluators was independently associated with overall image quality ratings. Compared with pediatric and adult cardiologists (reference group), non-cardiologist medical experts had perceived significantly higher overall quality scores (RR = 1.05, 95% CI [1.02, 1.08], p < .001). The mean perceived cardiac anomaly complexity score demonstrated a small inverse association with overall image quality; however, this relationship did not reach statistical significance (RR = 0.99, 95% CI [0.98, 1.00], p = 0.056).

### Correlations between evaluators’ perceptions of the four criteria of AI Cardiac Anomalies Images

Spearman’s rank-order correlations examine the relationships between physicians’ ratings of anatomical accuracy, text usefulness, image attractiveness, usability for medical education, and perceived cardiac anomaly complexity. Overall, the findings demonstrate strong and statistically significant positive associations among the core image-quality domains, alongside weak or negligible associations with perceived anomaly complexity (Table 3)

**Table 3:** Spearman’s Correlations Between the Medical Professionals Ratings of Cardiac Anomalies Ratings.

| **Parameter** | **Anatomic Accuracy** | **Text Usefulness** | **Image Attractiveness** | **Usability for education** |
| --- | --- | --- | --- | --- |
| Anatomic Accuracy of the images | 1 |  |  |  |
| Text usefulness for viewer | .699** |  |  |  |
| Image attractiveness | .590** | .637** |  |  |
| Usability for medical education | .829** | .784** | .644** |  |
| Perceived cardiac anomalies complexity score | -0.028 | -.051** | 0.009 | -.053** |
| **\*\*.** Correlation is significant at the 0.01 level (2-tailed). |  |  |  |  |

A strong positive correlation was observed between anatomical accuracy and usability for medical education (ρ = .829, p < .01), indicating that images perceived as anatomically accurate were substantially more likely to be judged as suitable for educational use. Similarly, text usefulness was strongly correlated with educational usability (ρ = .784, p < .01), underscoring the importance of clear and meaningful annotations in supporting the educational value of cardiac images. These findings suggest that both structural correctness and explanatory text play central roles in determining whether an image is perceived as pedagogically useful.

Moderate-to-strong positive correlations were also identified between anatomical accuracy and text usefulness (ρ = .699, p < .01), as well as between image attractiveness and both anatomical accuracy (ρ = .590, p < .01) and text usefulness (ρ = .637, p < .01). These associations indicate that visually appealing images were more likely to be perceived as accurate and well-labeled, although attractiveness alone was less strongly related to educational usability than accuracy and text clarity. In contrast, perceived cardiac anomaly complexity showed negligible or very weak correlations with most image-quality domains.

## Discussion

This study provides a detailed evaluation of how contemporary textl1ltol1limage systems perform when tasked with generating CHD illustrations for educational use. Several patterns emerged from our data. First, anatomical accuracy and educational usability of AIl1lgenerated images remained clearly inferior to the currently used human-modified illustrations, although Gemini NanoBanana consistently outperformed the ChatGPTl1lbased generators. Second, evaluators’ clinical roles influenced their judgments, with cardiology specialists consistently more critical than nonl1lcardiology physicians. Together, these findings place CHDl1lfocused image generation within a broader body of work showing that generative models can produce visually compelling medical imagery but still struggle to achieve the anatomical precision required for safe use in education.

The performance gradient between AIl1lgenerated outputs was particularly striking. The human-modified images achieved the highest proportions of anatomically accurate ratings, followed by Gemini NanoBanana, whereas both ChatGPTl1lderived systems were predominantly judged fabricated or incorrect. This is consistent with recently published work showing that Gemini produced the most anatomically correct representations of the human heart and outperformed other AI models such as DeepAI and Freepik [15]. Additionally, it is further supported by our previous work showing a large majority of ChatGPT’s DALL·E-3 images of common CHD lesions contained major inaccuracies in chamber relationships, outflow tract alignment, or great vessel branching [12]. Similar patterns have been documented in hand surgery and other anatomical domains, where AI generated images exhibit excellent detail -generated images exhibit excellent detail and apparent realism yet introduce fictitious or misplaced structures in almost all cases, with one study documenting an astonishing 99.8% of images containing fabricated anatomy [16]. In our multivariable model, image source remained the dominant predictor of overall quality even after adjusting for evaluator role and perceived lesion complexity, with both ChatGPT versions showing large reductions in adjusted quality and NanoBanana showing a smaller but significant decrement relative to the human-modified CHD Images. These observations indicate that text-to-image systems should not be treated as interchangeable; model choice has a substantial impact on anatomical precision, and even the comparatively stronger performers in this study did not match the standard set by expert curated-curated CHD illustrations [17].

Our analysis also underscores the central importance of labels and explanatory text in determining whether an image is educationally useful. The human-modified images achieved the highest proportion of ratings classifying labels as useful, while NanoBanana outperformed the ChatGPT-based generators, whose labels were frequently judged incorrect, ambiguous, or poorly aligned with the depicted structures.

Similar concerns have been raised in systematic evaluations of AI-generated anatomy and procedural images, where mislabeling, incomplete nomenclature, and inconsistent arrow placement undermine otherwise acceptable artwork [18]. From the perspective of cognitive load theory, such defects are not minor; unclear or inaccurate labeling increases extraneous cognitive load and diverts working memory resources away from learning, particularly in high-intrinsic load domains such as CHD [19]. In our correlation analysis, anatomical accuracy and label usefulness showed strong positive associations with educational usability, whereas attractiveness demonstrated weaker relationships with these constructs. This pattern suggests that structural accuracy and precise text are jointly necessary for pedagogical effectiveness and that aesthetics alone are poor predictors of educational value [20].based generators, whose labels were frequently judged incorrect, ambiguous, or poorly aligned with the depicted structures. Similar concerns have been raised in systematic evaluations generated anatomy and procedural images, where mislabeling, incomplete nomenclature, and inconsistent arrow placement undermine otherwise acceptable artwork intrinsic load domains such as CHD-based generators, whose labels were frequently judged incorrect, ambiguous, or poorly aligned with the depicted structures. Similar concerns have been raised in systematic evaluations -generated anatomy and procedural images, where mislabeling, incomplete nomenclature, and inconsistent arrow placement undermine otherwise acceptable artwork-intrinsic-load domains such as CHD.

AI-generated images are visually preferred but harbor non-trivial anatomical fabrications The dissociation between visual attractiveness and educational adequacy is a further notable finding. Gemini NanoBanana images were most frequently rated as attractive and surpassed even the human-modified images in this dimension, yet their anatomical correctness and label quality remained clearly inferior to the human-modified CHD Images, and their “as is” usability was much lower. Similar dissociations have been reported in CHD and general anatomy studies, where highly realistic AIl1lgenerated images are visually preferred but harbor nonl1ltrivial anatomical fabrications [12,21]. This creates a specific risk for CHD education. Learners must construct accurate three-dimensional understandings of atrial-ventricular relations, great artery positions, septal defects, and surgical pathways from d-dimensional understandings of atrial–ventricular relations, great artery positions, septal defects, and surgical pathways from two-dimensional images; visually polished but inaccurate depictions may be particularly likely to anchor incorrect mental representations that are difficult to unlearn. This creates a specific risk for CHD education. Visually polished but inaccurate depictions may be particularly likely to anchor incorrect mental representations that are difficult to unlearn [22–24]. The moderate correlations observed between attractiveness and educational usability in our study reinforce that aesthetic quality is, at best, a secondary consideration and cannot substitute for rigorous scrutiny of structural and textual accuracy.

The usability data emphasizes the need for expert-led adaptation of both the human-modified and AI-led adaptation of both reference and AI-generated images. The human-modified CHD Images were most often rated as usable “as is;” yet they still required modification in the majority of instances, whereas Gemini’s NanoBanana images were most frequently classified as usable only after modification, and ChatGPT-generated images were overwhelmingly deemed unsuitable for educational use. This pattern is consistent with experience from generated images were overwhelmingly deemed unsuitable for educational use. This pattern is consistent with experience from three-generated images were overwhelmingly deemed unsuitable for educational use. This pattern is consistent with experience from three-dimensional printed cardiac models and virtual reality or dimensional printed cardiac models and virtual reality or mixed-dimensional printed cardiac models and virtual reality or mixed-reality platforms, where raw outputs seldom meet instructional needs without targeted adjustments in orientation, segmentation, and annotation [23,25,26]. In routine practice, CHD teaching relies heavily on three dimensional and extended-dimensional and extended-reality representations that can be interactively manipulated to align with specific learning goals [27]. Educators and clinicians routinely rotate cardiac models, apply multiplanar cuts, adjust viewing angles, and adjust the visibility of selected regions in order to expose non-standard surgical perspectives and clarify complex intracardiac relationships [27]. In parallel, labels and overlays are tailored to draw attention to key physiologic or surgical features, and dynamic annotations are used to explain lesion-specific blood-flow patterns in virtual reality applications designed for complex congenital heart defects [28]. This type of deliberate manipulation, where educators highlight, suppress, and relabel structures to match the concept being taught, mirrors the substantial expert effort required to adapt AI-generated CHD images before they can function as safe and effective educational tools. Within this context, the high proportion of AI-generated images classified as usable only after modification reflects substantial remedial effort: restoring anatomical correctness, standardizing terminology, clarifying spatial orientation, and aligning each illustration with the cognitive demands of the target learner group. generated images classified as usable only after modification reflects substantial remedial effort: restoring anatomical correctness, standardizing terminology, clarifying spatial orientation, and aligning each illustration with the cognitive demands of the target learner group.-generated images classified as usable only after modification reflects substantial remedial effort: restoring anatomical correctness, standardizing terminology, clarifying spatial orientation, and aligning each illustration with the cognitive demands of the target learner group. These findings argue against the uncritical, routine, unsupervised use of AI -generated CHD imagery and instead support a workflow in which such images are treated as editable drafts subject to systematic expert revision [29].

The influence of clinical expertise on image appraisal provides an additional layer of insight. Cardiology specialists and pediatric consultants consistently assigned lower overall quality scores and higher complexity ratings than non-cardiology physicians, while non-cardiology experts tended to rate images more favorably. This pattern aligns with eye-tracking studies of medical image interpretation in cardiology, which show that experienced clinicians and less experienced readers both fixate on diagnostically critical regions but differ in how they interpret visual information and in the standards they apply when deciding whether an image is adequate [12,30]. In other evaluations of AI-generated CHD images, a similar pattern was observed, with nurses and trainees rating these images more favorably, while cardiology experts being more likely to detect inaccuracies and to question their suitability for teaching. In our study, the trend toward lower overall quality ratings as perceived anomaly complexity increased suggests that images representing more complex lesions, such as pulmonary atresia with ventricular septal defect and major aortopulmonary collaterals, or double-outlet right ventricle, were particularly likely to fall short of expert expectations. These are precisely the entities in which subtle distortions in spatial arrangement or great-vessel relationships can have major consequences for understanding physiology and management, reinforcing the need for heightened caution when using AI-generated images to illustrate high-complexity CHD [11].

When considered together, these results position text-to-image systems as promising but clearly bounded tools in CHD education and communication. Current general-purpose generators cannot replace curated CHD illustrations for high-stakes tasks such as core lesion teaching, examination preparation, or detailed pre-procedural explanation. However, the relatively stronger performance of Gemini NanoBanana compared with the ChatGPT-based generators, combined with the robust association between structural accuracy, label quality, and educational usability, suggests that AI-generated images may have meaningful roles when embedded in appropriately governed workflows. Potential applications include rapid prototyping of lesion-specific views, generating alternative angles or simplified versions of complex images tailored to different learner levels, and creating draft materials for patient and family counseling that are then verified and refined by CHD specialists [31,32]. To realize these benefits safely, institutions will need explicit policies that require expert review of AI-generated images, clarify when and how such images may be used, and promote transparency to learners regarding which materials are AI-derived. In this expert-supervised workflow, generative AI does not replace the expertise of cardiologists and medical illustrators; instead, it serves as a flexible visual engine that can accelerate and diversify image creation, while final responsibility for anatomical and educational integrity remains firmly with domain experts [33].

## Strengths and Limitations

This study has several strengths that enhance the credibility and relevance of its findings. The assessor-blinded design, with the human-modified images visually harmonized with AI outputs, minimized recognition bias and supported fair comparison across image sources. The inclusion of both normal heart anatomy and a broad spectrum of common CHDs, ranging from simple shunt lesions to highly complex single ventricle and outflow tract anomalies, allowed performance to be evaluated across clinically meaningful levels of complexity, which is often lacking in prior generative AI image studies that focus on limited or simplified anatomy. The use of a structured, multidomain scoring framework and mixed effects modeling further strengthened internal validity by capturing multiple dimensions of image quality and accounting for evaluator heterogeneity.

However, several limitations should be acknowledged. First, the study evaluated a fixed set of prompt structures and model configurations; different prompts, fine-tuning strategies, or future model updates might yield different performance, so the findings should not be generalized to all potential uses of these or other AI platforms before further research. Second, evaluators were limited to 20 healthcare, thus reducing generalizability to learners, allied health professionals, or educators in other settings with different expectations and visual literacy; therefore, more research is warranted. Third, although the rating tool captured key educational attributes, it did not directly assess downstream learning outcomes, such as diagnostic accuracy or long-term retention, which will be essential to determine real-world educational impact. Finally, because only a limited set of generators was examined, the study cannot address the rapidly expanding ecosystem of domain-specific or institutionally fine-tuned models that may perform differently for CHD anatomy.

## Conclusion

This blinded comparative study demonstrates that contemporary general-purpose text-to-image generators produce CHD illustrations that fall substantially short of curated educational standards across dimensions of anatomical accuracy, labeling quality, and educational utility. Gemini’s NanoBanana exhibited the strongest performance among tested GenAI systems, approaching but not equaling the human-modified reference images, while both ChatGPT-based platforms showed markedly diminished CHD quality, with degradation particularly pronounced for complex anatomical scenarios. Image source consistently emerged as the dominant predictor of overall quality. Moreover, the finding that cardiology specialists applied significantly more rigorous evaluation criteria than non-cardiology professionals underscores both the value of domain expertise in identifying structural errors and the vulnerability to misplaced confidence in visually persuasive but anatomically flawed outputs. Collectively, these results establish that AI-generated CHD imagery should function exclusively as expert-reviewed supplementary tools, suitable for rapid prototyping or teaching variants, rather than as independent educational resources. Our findings underscore the importance of robust institutional governance frameworks that define requirements for transparency, quality-assurance procedures, and careful selection of AI models, in addition to highlighting that systematic, evidence-based validation must precede any wider curricular integration of generative tools.

## Data Availability

All data produced in the present study are available upon reasonable request to the corresponding author

## Acknowledgement

While preparing this manuscript, the authors used ChatGPT-5 to refine the language and grammar, then the authors reviewed and edited the content as needed and they take full responsibility for the content of the submitted manuscript. The authors are grateful to the Deanship of Scientific Research, King Saud University, for funding through the Vice Deanship of Scientific Research Chairs. We also thank Hodhodata.com for their support in statistical analysis.

---

**Supplementary Table:**

| Variable | Clinical Role of Raters |  |  | p-value |
| --- | --- | --- | --- | --- |
|  | Cardiology Experts | Pediatrics Consultants | Non-cardiology medical experts |  |
| Anatomical accuracy of image |  |  |  |  |
| Accurate | 134 (19.9) | 97 (14.4) | 307 (15.2) | <0.001 |
| Midway | 118 (17.6) | 138 (20.5) | 539 (26.7) |  |
| Inaccurate | 420 (62.5) | 437 (65) | 1170 (58) |  |
| Usefulness of text for the viewer |  |  |  |  |
| Useful | 108 (16.1) | 113 (16.8) | 274 (13.6) | <0.001 |
| Midway | 128 (19) | 136 (20.2) | 680 (33.7) |  |
| Not useful | 436 (64.9) | 423 (62.9) | 1062 (52.7) |  |
| Attractivity of image for viewer |  |  |  |  |
| Attractive | 126 (18.8) | 112 (16.7) | 323 (16) | <0.001 |
| Midway | 125 (18.6) | 148 (22) | 762 (37.8) |  |
| Not attractive | 421 (62.6) | 412 (61.3) | 931 (46.2) |  |
| Usability of image for medical education |  |  |  |  |
| As is | 87 (12.9) | 63 (9.4) | 184 (9.1) | <0.001 |
| after modification | 160 (23.8) | 133 (19.8) | 540 (26.8) |  |
| Not useful for use | 425 (63.2) | 476 (70.8) | 1292 (64.1) |  |
| Overall quality of image rating, median (IQR) | 4 (4) | 4 (4) | 5 (4) | <0.001 |
| Perceived cardiac anomaly complexity, median (IQR) | 3 (1.25) | 2.38 (1.5) | 2.75 (1.25) | 0.001 |

